# Detection of *Mansonella ozzardi* in patients with acute febrile illness in Colombia

**DOI:** 10.1101/2024.02.24.24302413

**Authors:** Karl A. Ciuoderis, Mostafa Zamanian, Laura Perez, Claudia Patiño, Maria Angelica Maya, John D. Chan, Gavin A. Cloherty, Juan P. Hernandez-Ortiz, Jorge E. Osorio

**Affiliations:** Universidad Nacional de Colombia - UW-GHI One Health Colombia, Medellín, Colombia; University of Wisconsin-Madison, Global Health Institute, Madison, Wisconsin, USA; University of Wisconsin-Madison, Department of Pathobiological Sciences, Madison, Wisconsin, USA; Division of infectious Diseases, San Vicente Fundación Hospital, Medellín, Colombia; Abbott Diagnostics, Abbott Park, Illinois, USA

**Keywords:** Mansonella, Fever, Vector-borne disease, Colombia, Amazon

## Abstract

Mansonellosis is a vector-borne filariasis caused by parasitic nematodes of the genus *Mansonella*. The prevalence and health impact of mansonellosis is largely unknown, and there are no control programmes targeting this neglected tropical disease. Mansonellosis is prevalent in certain regions of Colombia, and while infection is often thought to be asymptomatic it may be associated with underrecognized clinical manifestations. In this study, we analyzed biobanked specimens from 905 patients participating in a febrile syndrome surveillance program in Colombia for evidence of *Mansonella ozzardi* infection, identifying four confirmed cases. While there have been prior reports on the incidence of mansonellosis in Colombia, this is the first report to our knowledge describing *M. ozzardi* microfilariae occurring in febrile patients in the country. Additional studies are needed to better understand the clinical consequences of mansonellosis within the complex tropical environment of Colombia, which is endemic for numerous other blood-transmitted and other vector-borne pathogens.

## INTRODUCTION

Mansonellosis is a tropical vector-borne neglected disease caused by filarial nematodes of the genus *Mansonella*. Two species of *Mansonella* have been described in Latin America; *M. ozzardi* has been widely detected in South and Central America, while *M. perstans* has been reported in the Amazon regions of Venezuela, Colombia, and Guyana^1^. Mansonellosis may be spread through infected biting midges (genus *Culicoides*) or blackflies (genus *Simulium*)^2^. Some areas in Colombia have been classified as endemic for *M. ozzardi*^*3*^, but very few reports have examined the incidence of this disease throughout the country^4,5^. However, a recent surveillance study conducted in the Colombian Amazon Basin found a high prevalence of *M. ozzardi* infection in this region^6^.

## RESULTS

Biobanked specimens obtained from 905 subjects (between 10 - 83 years old; 62.3% female and 37.7% male) in a Colombian health facility-based fever surveillance program were used for this study. These specimens were from two distinct locations in Colombia - Leticia, which is located in the Colombian Amazon River basin, and Villavicencio, in Central Colombia at the boundary of the Andes Mountains and the Eastern Plains (Figure 1A). At each location, patients were initially tested for common endemic pathogens (e.g. dengue virus, plasmodium parasites) using rapid diagnostic tests, and samples were shipped to a central laboratory in Medellín for more in depth analysis (Figure 1B). Over the course of this testing, microfilaria were incidentally observed in blood smears of four patients (Figure 1C). Infection by *M. ozzardi* was confirmed in each of these samples by PCR. Patients are referred to here by sample IDs not known to anyone outside the research group.

**Figure 1.**
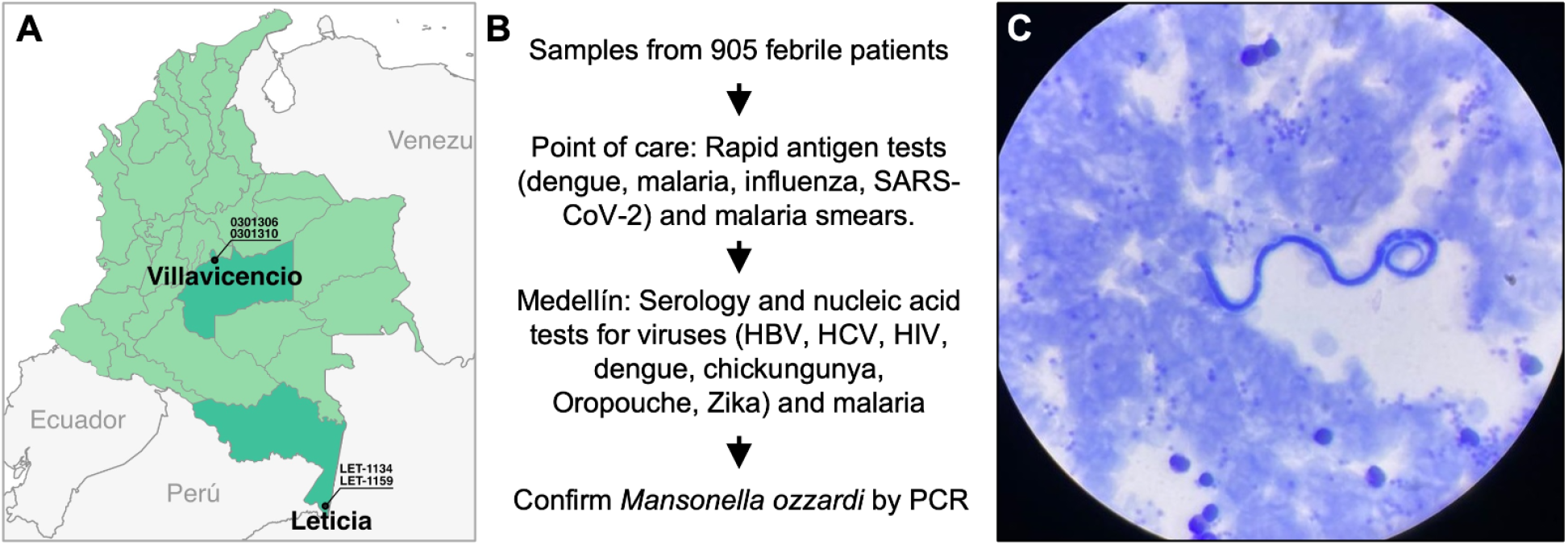
Detection of *Mansonella ozzardi* in biobanked samples from febrile patients. **(A)** Map showing the source of the biobanked specimens. Leticia is in the Southernmost part of the country (in the Amazon), while Villavicencio is in the center of the country (at the boundary of the Andes Mountains and Eastern Plains). *M. ozzardi* positive sample IDs are labeled at each location. **(B)** Workflow resulting in detection of mansonella from blood samples. A total of 905 patient samples were analyzed for common causes of febrile illness. Rapid antigen tests were performed at the site of sample collection (either Leticia or Villavicencio) to test for common infections that cause febrile illness, and blood smears for detection of malaria parasites were also prepared. Samples were then shipped to the central lab in Medellín, where more exhaustive testing was conducted using serological and nucleic acid tests. Blood smears were also interpreted by microscopy, and samples from four febrile patients were incidentally found to contain microfilariae that were later confirmed as *M. ozzardi* by PCR. **(C)** Giemsa-stained thick blood smear showing *M. ozzardi* microfilariae (100x magnification).

The samples from Villavicencio were from one indigenous and one non-indigenous patient, who both presented during November, 2021. Patient (ID 0301306) presented and self-reported symptoms including fever (body temperature ≥37.5°C two days before enrollment), fatigue, myalgia, headache, retro-orbital pain, vomit, dizziness, and shaking chills, and patient ID 0301310 similarly self-reported fever (body temperature ≥37.5°C three days before enrollment), fatigue, myalgia, headache, retro-orbital pain, vomit, dizziness, and shaking chills.

The positive samples in Leticia were both from indigenous Colombians living in the rural Mojoca-Jagua community who presented to the fever clinic between October 2022 - February 2023. Patient ID LET-1134 self-reported symptoms including fever (body temperature of 37.6°C at enrollment), fatigue, myalgia, headache, abdominal pain, nausea and dizziness. The second patient, ID LET-1259, presented with self-reported fever (body temperature of 37.9°C at enrollment), fatigue, myalgia, headache, retro-orbital pain, abdominal pain, joint pain, dizziness, and diarrhea.

Dengue fever was the presumptive clinical diagnosis for all patients. None of the patients reported traveling outside of the living area a month prior to sample collection. Additional characteristics of the mansonella cases are detailed in the Supplementary File (Table S1). Specimens (whole blood, serum, thick & thin blood smears, and nasopharyngeal swabs) were initially tested at the point of care using rapid diagnostic testing (RDT) and transported to a central laboratory in Medellín for extensive additional testing (RT-PCR, PCR, and microscopy) for various pathogens that could be the cause of febrile illness (Table S1) including arboviruses (dengue virus, Zika virus, chickungunya virus), Oropouche virus, *Plasmodium* parasites, hepatitis B (HBV) and C (HCV) viruses, human immunodeficiency virus (HIV), SARS-CoV-2 and influenza type A, B and A(H1N1). Laboratory testing found that samples from two patients (LET-1134 from Leticia, 0301310 from Villavicencio) were negative for all potential causes of febrile illness except for *Mansonella*. One sample from Villavicencio (0301306) was seropositive to HCV, and one sample from Leticia (LET-1259) had a dengue virus infection confirmed by RT-PCR and RDT (NS1 antigen). Details on laboratory testing and methods are provided in the supplementary file (Table S1 and S2). Microscopy of Giemsa-stained blood smears testing for malaria revealed the presence of microfilariae nematodes in samples from each patient (Figure 1C). PCR testing was conducted, confirming positive results for *M. ozzardi*. Patients were not subjected to treatment for mansonellosis, as there is no standard recommended treatment protocol. There was no patient follow-up to determine if symptoms persisted over time.

## DISCUSSION

This is one of the few studies describing cases of mansonellosis with active symptoms and clinical manifestations. Each of the four cases of mansonellosis were found incidentally during testing for other causes of febrile illness, whenever microfilariae were observed in blood smears looking for malaria parasites. The cases we have identified are an underestimation of the actual prevalence of mansonellosis in the country.

A review of the literature to search for cases of patients with mansonellosis in Colombia (PubMed search using the terms “mansonella” or “filaria” or “filarial nematode” in combination with “fever” or “Colombia”) shows no reports of mansonellosis after the 1980s, with the exception of our recent study published in 2023^6^. In the 1980’s a survey of 16 indigenous communities in the Colombian Amazon found 47.1% of samples were positive for *M. ozzardi*^*3*^. Prevalence of *M. ozzardi* was also estimated in several other regions of Colombia at that time, and the incidence varied depending on the location and demographics of the population. A high percentage (49%) of blood smears tested positive for *M. ozzardi* in mostly indigenous populations in the Southeast of the country^4^, while primarily caucasian locations had lower rates (between 2 - 4% positivity)^7^. There was also a recent incidental finding of *M. ozzardi* in an indigenous child suffering from T-cell lymphoma^8^. None of these studies addressed whether *M. ozzardi* infection was linked to clinical manifestations of illness.

While the majority of mansonella affected individuals are reported to be asymptomatic and the infection is often described as self-limiting, it is also appreciated that the disease may not be entirely non-pathogenic^2,9^. Whenever infected patients do present with clinical manifestations, these commonly include symptoms related to the site of infection (e.g.itching, ocular lesions^10,11^) or the host immune response to parasite antigens (e.g. eosinophilia^12^). The latter may also underlie febrile symptoms. Circulating immune complexes are thought to induce fever and inflammatory responses in other filarial infections^13,14^, and these complexes are also present in patients infected with *Mansonella*^*15*^.

Chronic *Mansonella* infections may also affect host immune responses to pathogens that are co-endemic in many regions in South America and Africa. It has been suggested that the immune responses associated with severe malaria were reduced in individuals co-infected with *M. perstans*^*16*^. Filariasis was associated with an impaired T cell response to malaria antigens in regions co-endemic for *Plasmodium falciparum* and the filaria *M. perstans* and *Wuchereria bancrofti*^*17*^. Infection with the filarial parasite that causes onchocerciasis is associated with decreased immunity to mycobacteria^17,18^. Mansonellosis may also impact the endemicity of other vector-borne diseases such as dengue. Microfilarial enhancement of arboviral transmission has been shown for other parasites (e.g. *Brugia* and *Onchocerca* species), and *M. ozzardi* microfiliariae have been shown to be capable of penetrating the midgut of *Aedes aegypti* mosquitoes^19^. Consequently, there is a need for further evidence-based information regarding the health impact of mansonellosis. While this parasite is endemic in many tropical regions, the burden of this disease is poorly understood and there are no control programs in place^20^.

## MATERIALS AND METHODS

Study area and population: A biobank of repository specimens was obtained from a healthcare facility-based fever surveillance program involving 2,977 subjects, which was conducted from December 2020 to February 2023 in two regions of Colombia (Leticia and Villavicencio). During informed consent, these subjects agreed and understood that data and specimens that were initially provided for the fever surveillance program would be used not only for that program for which they were collected, but also for other future studies after patient data were anonymized. A total of 905 (30.4%) subjects were randomly selected from the biobank of 2,977 for analysis in this study. Patients are referred to by sample IDs not known to anyone outside the research group.

Sample collection and point of care testing: Venous whole blood sample was collected from each patient and tested by point of care rapid diagnostic tests for dengue (SD Bioline Dengue Duo, Abbott Labs), malaria (SD Bioline Malaria Ag P.f/Pan, Abbott Labs), influenza (SD Bioline Influenza, Abbott Labs), SARS-CoV-2 (Panbio COVID-19 Ag Rapid Test, Abbott Labs). An additional sample was collected in a serum separator tube, centrifuged, and serum was stored as 2 ml aliquots at −80°C.

Serological and nucleic acid testing: Samples were shipped to the One Health Genomic Lab at the Universidad Nacional in Medellin, Colombia for serological and nucleic acid tests. These include tests for hepatitis B and C (ARCHITECT HBsAG Qualitative II kit and ARCHITECT HCVcAG kit, Abbott Labs), HIV (ARCHITECT HIV Ag/Ab Combo, Abbott Labs) which were detected by chemiluminescence immunoassay using the Architect I1000 system, and the Arbovirus trioplex RT-PCR panel for dengue, zika and Chickungunya virus (Abbott Labs).

Giemsa stained thick and thin blood smears were also analyzed by microscopy for malaria diagnosis, during which time *Mansonella* microfilariae were incidentally observed. *M. ozzardi* diagnosis was confirmed using PCR Real time PCR test was carried out in a reaction volume of 50 μL using iTaq DNA polymerase kit (BioRad, Hercules, California) and run in a BioRad CFX96 thermal cycler (BioRad, Hercules, California). PCR conditions, primers and probe for detection of the Internal transcribed spacer 1 (ITS1) gene of the genus Mansonella spp. were followed as described in reference^21^.

## Data Availability

All data produced in the present work are contained in the manuscript.

## ABBREVIATIONS

RDT: Rapid diagnostic testing
RT-PCR: Reverse transcription polymerase chain reaction
PCR: Polymerase chain reaction
NS1: Non-structural protein 1
IgG: immunoglobulin G
IgM: immunoglobulin M
SD: Standard diagnostic
COVID-19: Coronavirus disease 2019
HBV: hepatitis B virus
HCV: hepatitis C virus
HIV: human immunodeficiency virus

## DECLARATIONS

### Ethics Approval and consent to participate

This work followed the EQUATOR Reporting Guidelines and conformed to the Declaration of Helsinki and to the international ethical guidelines for health-related studies involving humans, specifically indigenous people. The study protocol was reviewed and approved by the ethics committee of the Corporacion para Investigaciones Biologicas (CIB #10102022). Written informed consent (18 years and older) or assent (5 to 17 years) was initially obtained from each subject. Parents or legal guardians of minors provided written informed consent on their behalf. Written approval for specimen and data storage for use in future research studies was also provided. Only anonymized samples and data were used in this study.

### Availability of supporting data

The authors confirm that the data supporting the findings of this study are included in this published article. Deidentified case study data or additional information are available from the corresponding author upon request.

### Funding

Funding for this project was provided by Abbott Laboratories through the Abbott Pandemic Defense Coalition (APDC), the Universidad Nacional de Colombia (UNC), and the University of Wisconsin-Madison-Global Health Institute (UW-GHI).

## Acknowledgments

Authors would like to thank the E.S.E Hospital San Rafael de Leticia, the Laboratorio Departamental de Salud Pública de Leticia, and the staff of One Health lab for their assistance to this study. We sincerely thank Julian Rodriguez and Corporacion Corpotropica for their assistance to this work. Finally, thanks to the Abbott Pandemic Defense Coalition for the provision of RDT kits. This work was conducted as part of the Project “Caracterización epidemiológica de casos asociados a síndrome febril indiferenciado en áreas de alto riesgo de transmisión de enfermedades transmitidas por vectores”.

## Conflict of Interest Disclosures

GC receives salaries from and / or owns stock in Abbott. JO, JP and/or their institutions receive funding from Abbott Diagnostics.

## Author Contributions

Concept and design: KC, MZ, JO, JPHO; Writing: all authors; Formal Analysis: all authors; Drafting and critical revision of the manuscript for important intellectual content: all authors. Sample processing and data analysis: KC, LP, CP; Funding acquisition: MZ, JO, JPHO; Writing – review & editing: All authors.

## Notes

### Author Declarations

Ethics committee of the the Corporacion para Investigaciones Biologicas gave ethical approval for this work.

## REFERENCES

1. Ta-Tang T-H, Luz SLB, Merino FJ, de Fuentes I, López-Vélez R, Almeida TAP, Lanza M, Abrahim CMM, Rubio JM., 2016. Atypical Mansonella ozzardi Microfilariae from an Endemic Area of Brazilian Amazonia. Am J Trop Med Hyg 95: 629–632

2. Lima NF, Veggiani Aybar CA, Dantur Juri MJ, Ferreira MU., 2016. Mansonella ozzardi: a neglected New World filarial nematode. Pathog Glob Health 110: 97–107

3. Kozek WJ, D’Alessandro A, Silva J, Navarette SN., 1982. Filariasis in Colombia: prevalence of mansonellosis in the teenage and adult population of the Colombian bank of the Amazon, Comisaria del Amazonas. Am J Trop Med Hyg 31: 1131–1136

4. Lightner LK, Ewert A, Corredor A, Sabogal E., 1980. A parasitologic survey for Mansonella ozzardi in the Comisaría del Vaupés, Colombia. Am J Trop Med Hyg 29: 42–45

5. Kozek WJ, Palma G, Henao A, García H, Hoyos M., 1983. Filariasis in Colombia: prevalence and distribution of Mansonella ozzardi and Mansonella (=Dipetalonema) perstans infections in the Comisaría del Guainía. Am J Trop Med Hyg 32: 379–384

6. Dahmer KJ, Palma-Cuero M, Ciuoderis K, Patiño C, Roitman S, Li Z, Sinha A, Hite JL, Bellido Cuellar O, Hernandez-Ortiz JP, Osorio JE, Christensen BM, Carlow CKS, Zamanian M., 2023. Molecular surveillance detects high prevalence of the neglected parasite Mansonella ozzardi in the Colombian Amazon. J Infect Dis

7. Kozek WJ, Palma G, Valencia W, Montalvo C, Spain J., 1984. Filariasis in Colombia: prevalence of Mansonella ozzardi in the Departamento de Meta, Intendencia del Casanare, and Comisaría del Vichada. Am J Trop Med Hyg 33: 70–72

8. Suárez Mattos A, Calderón A, Acevedo A., 2018. Hallazgo incidental de microfilarias de Mansonella ozzardi en un paciente pediátrico con linfoma T anaplásico. Revista Colombiana de Cancerología 22: 88–91

9. Adami YL, Rodrigues G, Alves MC, Moraes MAP, Banic DM, Maia-Herzog M., 2014. New records of Mansonella ozzardi: a parasite that is spreading from the state of Amazonas to previously uninfected areas of the state of Acre in the Purus River region. Mem Inst Oswaldo Cruz 109: 87–92

10. Bartoloni A, Cancrini G, Bartalesi F, Marcolin D, Roselli M, Arce CC, Hall AJ., 1999. Mansonella ozzardi infection in Bolivia: prevalence and clinical associations in the Chaco region. Am J Trop Med Hyg 61: 830–833

11. Vianna LMM, Martins M, Cohen MJ, Cohen JM, Belfort R Jr., 2012. Mansonella ozzardi corneal lesions in the Amazon: a cross-sectional study. BMJ Open 2

12. Hochberg NS, Dinculescu VV, Nutman TB., 2023. Case 17-2023: A 58-Year-Old Woman with Fatigue, Abdominal Bloating, and Eosinophilia. N Engl J Med 388: 2180–2189

13. Kar SK, Mania J, Kar PK., 1993. Humoral immune response during filarial fever in Bancroftian filariasis. Trans R Soc Trop Med Hyg 87: 230–233

14. Senbagavalli P, Anuradha R, Ramanathan VD, Kumaraswami V, Nutman TB, Babu S., 2011. Heightened measures of immune complex and complement function and immune complex-mediated granulocyte activation in human lymphatic filariasis. Am J Trop Med Hyg 85: 89–96

15. Godoy GA., 1998. Circulating immune complexes in Mansonella ozzardi infection. Ann Trop Med Parasitol 92: 895–896

16. Dolo H, Coulibaly YI, Dembele B, Konate S, Coulibaly SY, Doumbia SS, Diallo AA, Soumaoro L, Coulibaly ME, Diakite SAS, Guindo A, Fay MP, Metenou S, Nutman TB, Klion AD., 2012. Filariasis attenuates anemia and proinflammatory responses associated with clinical malaria: a matched prospective study in children and young adults. PLoS Negl Trop Dis 6: e1890

17. Metenou S, Dembele B, Konate S, Dolo H, Coulibaly YI, Diallo AA, Soumaoro L, Coulibaly ME, Coulibaly SY, Sanogo D, Doumbia SS, Traoré SF, Mahanty S, Klion A, Nutman TB., 2011. Filarial infection suppresses malaria-specific multifunctional Th1 and Th17 responses in malaria and filarial coinfections. J Immunol 186: 4725–4733

18. Metenou S, Babu S, Nutman TB., 2012. Impact of filarial infections on coincident intracellular pathogens: Mycobacterium tuberculosis and Plasmodium falciparum. Curr Opin HIV AIDS 7: 231–238

19. Vaughan JA, Bell JA, Turell MJ, Chadee DD., 2007. Passage of ingested Mansonella ozzardi (Spirurida: Onchocercidae) microfilariae through the midgut of Aedes aegypti (Diptera: Culicidae). J Med Entomol 44: 111–116

20. Mediannikov O, Ranque S., 2018. Mansonellosis, the most neglected human filariasis. New Microbes New Infect 26: S19–S22

21. Bassene H, Sambou M, Fenollar F, Clarke S, Djiba S, Mourembou G L Y AB, Raoult D, Mediannikov O., 2015. High Prevalence of Mansonella perstans Filariasis in Rural Senegal. Am J Trop Med Hyg 93: 601–606

