## Supplementary File 1 for "Detection of *Mansonella ozzardi* in patients with acute febrile illness in Colombia"

**SUPPLEMENTARY MATERIAL**

| Testing conducted | Reference method | Patient Samples | | | |
|---|---|---|---|---|---|
| | | LET-1134 | LET-1259 | 0301306 | 0301310 |
| SARS-CoV-2 RDT | Panbio COVID-19 Ag Rapid Test, Abbott, Illinois, USA | Negative | Negative | Negative | Negative |
| Influenza RDT | SD Bioline Influenza, Abbott, Illinois, USA | Negative | Negative | Negative | Negative |
| Dengue virus RDT | Bioline Dengue Duo (Dengue NS1 Ag + IgG/IgM), Abbott, Illinois, USA. | Negative | Positive | Negative | Negative |
| Malaria RDT | SD Bioline Malaria Ag P.f/Pan, Abbott, Illinois, USA | Negative | Negative | Negative | Negative |
| HBV | ARCHITECT HBsAG Qualitative II kit, Abbott, Illinois, USA | Negative | Negative | Positive | Negative |
| HCV | ARCHITECT HCVcAG, Abbott, Illinois, USA | Negative | Negative | Positive | Negative |
| HIV | ARCHITECT HIV Ag/Ab Combo, Abbott, Illinois, USA | Negative | Negative | Negative | Negative |
| Arbovirus trioplex RT-PCR (Dengue virus, Zika virus, Chickungunya virus) | Waggoner, et al. Emerg Infec Dis. 2016 22(7):1295-1297. | Negative | Positive | Negative | Negative |
| Oropouche virus | Ciuoderis, et al. Emerg. Microb & Infec. 2022 11:1, 2645-2657. | Negative | Negative | Negative | Negative |
| Malaria PCR | Kamau, et al.. PLoS One. 2013 29;8(8):e71539. | Negative | Negative | Negative | Negative |
| Malaria Microscopy | Opoku, et al. Malar J. 2023 4;22(1):76. | Negative | Negative | Negative | Negative |
| Mansonella Microscopy | Dahmer et al. J infect dis, 2023, 228 (10):1441-1451. | Positive | Positive | Positive | Positive |
| Mansonella PCR | Bassene et al. Am J Trop Med Hyg. 2015;93(3):601-6. | Positive | Positive | Positive | Positive |

**Supplementary Table S1. Laboratory testing conducted in the four mansonellosis cases found in this study.** Abbreviations: RDT: rapid diagnostic testing; RT: reverse transcription: PCR: polymerase chain reaction; HBV: hepatitis B virus; HCV: hepatitis C virus; HIV: human immunodeficiency virus; HBsAG: hepatitis B virus surface antigen; HCVcAG: hepatitis C virus core antigen; SARS-COV-2: Severe acute respiratory syndrome coronavirus 2.

| Location | Mansonella PCR | Acute infection | | | | Chronic infection | | |
|---|---|---|---|---|---|---|---|---|
| | | Dengue | Malaria | Other * | Unknown etiology | HCV | HIV | HBV |
| Villavicencio | Positive (n=2) | 0 | 0 | 0 | 2 | 0 | 0 | 0 |
| | Negative (n=598) | 20 | 13 | 9 | 556 | 0 | 9 | 5 |
| Leticia | Positive (n=2) | 1 | 0 | 0 | 1 | 0 | 0 | 0 |
| | Negative (n=303) | 56 | 9 | 24 | 216 | 0 | 6 | 6 |

**Supplementary Table S2. Laboratory testing results obtained from 905 repository specimens from febrile patients.** Other* includes pathogens such as influenza virus, Severe acute respiratory syndrome coronavirus 2, *Leptospira* spp, Zika virus, Chikungunya virus, Oropouche virus.
